# Analysis of clinical trial registry entry histories using the novel R package cthist

**DOI:** 10.1101/2022.01.20.22269538

**Authors:** Benjamin Gregory Carlisle

**Affiliations:** Berlin Institute of Health at Charité – Universitätsmedizin Berlin, QUEST Center for Responsible Research, Charitéplatz 1, 10117 Berlin, Germany

## Abstract

Historical clinical trial registry data can only be retrieved by manually accessing individual clinical trials through registry websites. This limits the feasibility, accuracy and reproducibility of certain kinds of research on clinical trial activity and presents challenges to the transparency of the enterprise of human research. This paper presents `cthist`, a novel, free and open source *R* package that enables mass downloading of clinical trial registry entry histories and returns structured data for analysis.

Documentation of the implementation of the package `cthist` is provided, as well as 3 brief case studies with example code.

## Introduction

Prospective registration of clinical trials in a public database such as ClinicalTrials.gov or the German Clinical Trials Register, DRKS.de is ethically required of investigators by the Declaration of Helsinki,^1^ a prerequisite for publication according to the ICMJE,^2^ and for certain clinical trials, it is mandated by law.^3^ Preregistration of a clinical trial helps to reduce certain biases in the medical literature, such as selective reporting and non-publication bias. Reviewers of clinical trial journal publications can use registry entries to ensure that the published record corresponds to the trial that was planned. Clinical trial registries are also used as tools to help enrol prospective patients, providing them with information about new and ongoing studies by location and disease area. Systematic reviewers make use of clinical trial registries to synthesize available clinical evidence. Meta-researchers analyze data from trial registries to describe programmes of human research, evaluate them, or hold clinical investigators accountable to common standards of good research practice.

Clinical trial registry entries on ClinicalTrials.gov can be modified by the responsible party at any time, and changes to clinical trial registry entries between initial registration and last registration are common.^4^ Researchers who analyze clinical trial registry data but fail to account for potential changes to registry entries run the risk of making serious methodological errors, such as failure to account for variable follow-up time to an event of interest.^5^ Certain research questions and insights into the enterprise of human research are not possible or not feasible to address without a means of accessing and analyzing the history of changes for an entire cohort of clinical trials in a systematic way.

Prior to the publication of this package, options for accessing historical trial registry data were limited. There is no API access to historical clinical trial registry data for ClinicalTrials.gov or DRKS.de. Hence, the most straightforward way to access historical versions of a trial registry entry was by manually visiting the website for the clinical trial registry and recording the trial data systematically in a spreadsheet. While this was often feasible when considering a single clinical trial’s history, this method applied to a large cohort of clinical trials is extremely labour-intensive, error-prone and limits reproducibility. The challenge this presents to the feasibility of certain kinds of meta-research has been remarked on elsewhere. (See Al-Durra et al 2020,^6^ supplementary appendix L.)

Alternatively, a researcher could download the entire database of a clinical trial registry at regular intervals; however this is extremely resource-intensive, both in terms of data storage and the time required to process the data, which also limits the reproducibility of these methods.

In order to provide a tool that makes clinical trial registry history research accessible, feasible and reproducible for patients, reviewers, meta-researchers and systematic reviewers, `cthist`, the *R* package presented here, was developed. This package provides functions that allow access to historical clinical trial registry data from ClinicalTrials.gov and DRKS.de without bringing about the human error that would result from manual searches and extraction, or the resources that would be required for regularly mass-downloading and processing of the entire registry database. In what follows, the implementation of `cthist` is presented and its use is described with 3 potential case studies with example code.

## Methods

### Availability and requirements

The R package `cthist` can be downloaded from CRAN,^7^ or the development version can be installed from GitHub (https://github.com/bgcarlisle/cthist). Internal package documentation provides arguments and examples for the included functions. The package was written for *R*^8^, and depends on the following *R* packages: `dplyr`,^9^ `httr`,^10^ `jsonlite`,^11^ `magrittr`,^12^ `readr`,^13^ `rlang`,^14^ `rvest`,^15^ `selectr`,^16^ `stringr`,^17^ `tibble`.^18^

### Package implementation

The `cthist` package provides functions for downloading historical clinical trial registry data for trials registered on ClinicalTrials.gov and trials registered on DRKS.de. Future versions may include functions to download historical clinical trial registry data from other registries.

The functions for downloading historical trial data have been implemented for both ClinicalTrials.gov and DRKS.de. As much as possible given the constraints of the source database, the output returned by the function for ClinicalTrials.gov is similar to the output returned by the corresponding function for DRKS.de.

Six functions are provided by `cthist`: `clinicaltrials_gov_dates()`, `drks_de_dates()`, `clinicaltrials_gov_version()`, `drks_de_version()`, `clinicaltrials_gov_download()` and `drks_de_download()`. Three allow downloading of data from ClinicalTrials.gov, and the other three are their counterparts for DRKS.de. These are described in detail below.

### clinicaltrials_gov_dates() and drks_de_dates()

The functions `clinicaltrials_gov_dates()` and `drks_de_dates()` take a trial registration number (NCT number or DRKS id, respectively) as an argument and provide a vector of ISO-8601 formatted dates, one for each update to the registry entry history. For ClinicalTrials.gov, these dates are retrieved by downloading the HTML for the index of history changes, selecting all the cells in the “Submitted Date” column from the table of study record versions, extracting the text using the `rvest` *R* package, and re-formatting the dates.

Similarly for DRKS.de, dates are retrieved by downloading the HTML for the change history page, selecting all the cells in the “Date” column from the published versions table, extracting the text using the `rvest` *R* package, and reformatting the dates.

These functions then return a vector of dates in the case of success, and in the case of an error (e.g. inability to connect to the internet), they return the word “Error” and print out an explanatory error message in the *R* console.

### clinicaltrials_gov_version()

The function `clinicaltrials_gov_version()` downloads clinical trial data for the NCT number and version number (starting with 1) indicated by the function’s arguments. The HTML for the historical version page is downloaded using `rvest` and individual data points are extracted using a combination of cascading style sheet (CSS) selectors and regular expressions (regex). A list of 14 data points are returned by this function: overall status, enrolment, start date, primary completion date, primary completion date type, minimum age, maximum age, sex, gender based, accepts healthy volunteers, inclusion criteria, outcome measures, contacts and sponsors.

The overall status for the version of the trial in question is drawn from cells in the table with CSS `id` attribute `#StudyStatusBody` that have text that matches the regular expression `Overall Status: ([A-Za-z,]+)`.

Enrolment is extracted from cells from the table with CSS `id` attribute `#StudyDesignBody` that have text that matches the regular expression `Enrollment: ([A-Za-z0-9 \\[\\]]+)`. This text includes both the enrolment number (e.g. “50”) and the enrolment type (“Anticipated” or “Actual”), which are split into separate columns by the `clinicaltrials_gov_download()` function.

The study start date is extracted from cells in the table with CSS `id` attribute `#StudyStatusBody` that have text that matches the regular expression `Study Start: ([A-Za-z0-9,]+)`. The text of the date is then transformed from its original format (Month Date, Year) to ISO-8601 (YYYY-MM-DD). In the case that start dates are given only as accurately as the month, the date is rounded to the beginning of the month (e.g. “September 2009” would be rounded to 2009-09-01).

The primary completion date is extracted from cells with the CSS `id` attribute `#StudyStatusBody` that have text that matches the regular expression `Primary Completion: ([A-Za-z0-9, \\[\\]]+)`. The text of the date is then transformed from its original format (Month Date, Year) to ISO-8601 (YYYY-MM-DD). In the case that start dates are given only as accurately as the month, the date is rounded to the beginning of the month (e.g. “September 2009” would be rounded to 2009-09-01).

The primary completion date type is extracted from the same table cell as the primary completion date, but regex filtering for text enclosed by square brackets (`\\[[A-Za-z]+\\]`).

The minimum age, maximum age, sex, gender based, and accepts healthy volunteers data are extracted from cells in the table with CSS `id` attribute `#EligibilityBody` with text that matches the following regexes: `Minimum Age: ([0-9]+) Years`, `Maximum Age: ([0-9]+) Years`, `Sex: ([A-Za-z]+)`, `Gender based: ([A-Za-z]+)`, `Accepts Healthy Volunteers: ([A-Za-z]+)`, respectively.

Inclusion criteria are extracted as the contents of the cell of the table with CSS `id` attribute `#EligibilityBody` that immediately follows the cell that contains the label “Criteria:”. The lines of text comprising the contents of this cell are encoded as JavaScript Object Notation (JSON) to preserve the data structure.

Outcome measures are extracted from rows of the table with CSS `id` attribute `#ProtocolOutcomeMeasuresBody`. Each row in the original HTML table that contains an outcome measure is copied to a data frame with three columns: `section`, `label` and `content`. Because section headings are encoded in the original HTML as table rows where the second cell contains no text, the `section` is the text of the most recently preceding row where the second cell is empty. The `label` is the text in the first cell of the row. The `content` is the text contained in the second cell of the row. The data frame is encoded as JSON to preserve the data structure.

Contact information is extracted from rows of the table with CSS `id` attribute `#ContactsLocationsBody` that come before a row labelled “Locations:”. Each row in the original HTML table that contains contact information is copied to a data frame with two columns: `label` and `content`. The `label` is the text in the first cell of the row. The `content` is the text contained in the second cell of the row. This data frame is encoded as JSON to preserve the data structure.

Sponsors and collaborators are extracted from rows of the table with CSS `id` attribute `#SponsorCollaboratorsBody`. Each row in the original HTML table that contains sponsor or collaborator information is copied to a data frame with two columns: `label` and `content`. The `label` is the text of the first cell of the row, and the `content` is the text of the second cell. This data frame is encoded as JSON to preserve the data structure.

Upon successful download and parsing of a clinical trial registry history entry, the function returns a list of these 14 data points. In case of error, the text “Error” is returned. See *Table 1* for a list of the extracted data points and the CSS selectors used to identify them on ClinicalTrials.gov, and the corresponding data points and CSS selectors from DRKS.de for comparison.

**Table 1.** Cascading Stylesheet (CSS) selectors indicating the HTML elements on ClinicalTrials.gov and DRKS.de from which each of the extracted data points are taken by `clinicaltrials_gov_version()` and `drks_de_version()`, respectively

| Data point | ClinicalTrials.gov | DRKS.de |
| --- | --- | --- |
| Overall status | #StudyStatusBody | - |
| Recruitment status | - | li.state |
| Enrolment | #StudyDesignBody | li.targetSize |
| Enrolment type | - | li.targetSize |
| Start date | #StudyStatusBody | li.schedule |
| Primary completion date | #StudyStatusBody | - |
| Primary completion date type | #StudyStatusBody | - |
| Closing date | - | li.deadline |
| Minimum age | #EligibilityBody | li.minAge |
| Maximum age | #EligibilityBody | li.maxAge |
| Sex | #EligibilityBody | - |
| Gender | - | li.gender |
| Gender based | #EligibilityBody | - |
| Accepts healthy volunteers | #EligibilityBody | - |
| Inclusion criteria | #EligibilityBody | - |
| Additional inclusion criteria | - | .inclusionAdd |
| Exclusion criteria | - | .exclusion |
| Primary outcomes | - | p.primaryEndpoint |
| Secondary outcomes | - | p.secondaryEndpoints |
| Outcome measures | #ProtocolOutcomeMeasuresBody | - |
| Contacts | #ContactsLocationsBody | ul.addresses li.address |
| Sponsors | #SponsorCollaboratorsBody | - |

### drks_de_version()

The function `drks_de_version()` downloads clinical trial data for the DRKS number and version number (starting at 1) indicated by the function’s arguments. The HTML for the historical version page is downloaded using `rvest` and individual data points are extracted using a combination of cascading style sheet (CSS) selectors and regular expressions (regex). A list of 13 data points are returned by this function: recruitment status, start date, closing date, enrolment, enrolment type, minimum age, maximum age, gender, additional inclusion criteria, exclusion criteria, primary outcomes, secondary outcomes and contacts.

Because the data provided in the DRKS.de historical version page do not have a one-to-one correspondence with their counterparts on ClinicalTrials.gov, the columns extracted from the two registries differ.

The recruitment status for the version of the trial in question is drawn from the bullet point with CSS selector `li.state`, taking the text that follows “Recruitment Status:”.

The start date is taken from the bullet point with CSS selector `li.schedule`, extracting text that matches the regex `[0-9]{4}/[0-9]{2}/[0-9]{2}` (a numeric date with format YYYY/MM/DD). The date is reformatted to ISO-8601 (YYYY-MM-DD).

The closing date is taken from the bullet point with CSS selector `li.deadline`, extracting text that matches the regex `[0-9]{4}/[0-9]{2}/[0-9]{2}` (a numeric date with format YYYY/MM/DD). The date is reformatted to ISO-8601 (YYYY-MM-DD).

Enrolment is taken from the bullet point with CSS selector `li.targetSize`, extracting only numeric text that matches the regex `[0-9]+`.

Enrolment type is taken from the bullet point with CSS selector `li.running`, extracting the text that follows “Planned/Actual:”.

Minimum age and maximum age are taken from the bullet points with CSS selectors `li.minAge`, `li.maxAge`, respectively extracting the text that follows “Minimum Age:” and “Maximum Age:”, respectively.

Gender is taken from the bullet point with CSS selector `li.gender`, extracting the text that follows “Gender:”.

Additional inclusion criteria and exclusion criteria are taken from text elements with CSS selectors `.inclusionAdd` and `.exclusion`, respectively, encoded as JSON to preserve the data structure.

Primary and secondary outcomes are taken from text elements with CSS selectors `p.primaryEndpoint` and `p.secondaryEndpoints`, respectively, encoded as JSON to preserve the data structure.

Contact information is returned as a data frame with columns `label`, `affiliation`, `telephone`, `fax`, `email`, `url`. Each row in this data frame is populated by one bullet with CSS selector `ul.addresses li.address` from the original HTML. The `label` column is extracted from the HTML `<label>` node. The `affiliation` column is extracted from the bullet with CSS selector `li.address-affiliation`. The `telephone`, `fax`, `email` and `url` columns are extracted from the bullets with CSS selector `.address-telephone`, `.address-fax`, `.address-email` and `.address-url`, respectively excluding the label text for each. The data frame is encoded as JSON to preserve the data structure.

Upon successful download and parsing of a clinical trial registry history entry, the function returns a list of these 13 data points. In case of error, the text “Error” is returned. See *Table 1* for a list of the extracted data points and the CSS selectors used to identify them on ClinicalTrials.gov, and the corresponding data points and CSS selectors from DRKS.de for comparison.

### clinicaltrials_gov_download() and drks_de_download()

The functions `clinicaltrials_gov_download()` and `drks_de_download()` loop through the trial registry numbers provided to them in the first argument (NCT numbers or DRKS id’s, respectively), and for each one, download a list of all the dates on which the trial registry entry was updated, using `clinicaltrials_gov_dates()` or `drks_de_dates()`, respectively (described above). For each version of each trial, the functions download the clinical trial registry entry version using `clinicaltrials_gov_version()` or `drks_de_version()`, respectively. Each downloaded registry entry historical version is written to the filename specified in the second argument to the function, formatted as a CSV.

These functions also implement automated error-checking on completion, in order to ensure the accurate retrieval of large sets of clinical trial registry entry histories. If any version of the clinical trials to be downloaded returned an error, these functions return FALSE, otherwise they return TRUE. These functions also implement automated checking for already-downloaded data when starting, to allow for re-starting partially completed downloads.

## Results

The following is a description of 3 case studies of research questions regarding clinical trial registry entries that are difficult or un-feasible to answer without a tool for mass-downloading clinical trial registry history data. Case studies 1 and 3 provide example code for analyzing clinical trials from ClinicalTrials.gov and example 2 provides example code for DRKS.de, however each of the three could be re-written easily for the other clinical trial registy with minor changes.

### Case study 1: assessing change in length to recruitment period

Changes in recruitment length to a clinical trial have been used as one part of a measure of the feasibility of clinical trials.^19^ In order to evaluate changes to recruitment period lengths for a cohort of clinical trials, it is necessary to mass-download enrolment data from historical versions of ClinicalTrials.gov records.

As described in *R Code Box 1*, clinical trials meeting the study’s inclusion criteria were downloaded by performing a search via the web front-end of ClinicalTrials.gov. Search results were saved as a comma-separated value (CSV) file named `SearchResults.csv`. The `NCT Number` column in this file is parsed and used as an argument for the `clinicaltrials_gov_download()` function implemented in package `cthist`, which then downloads the records for all the clinical trial registry entries in this sample and saves them as `historical_versions_1.csv`. This script parses these results and the percentage change between the first registry entry version posted with an overall status of “Recruiting” and the registry entry version that was active 1 year later.

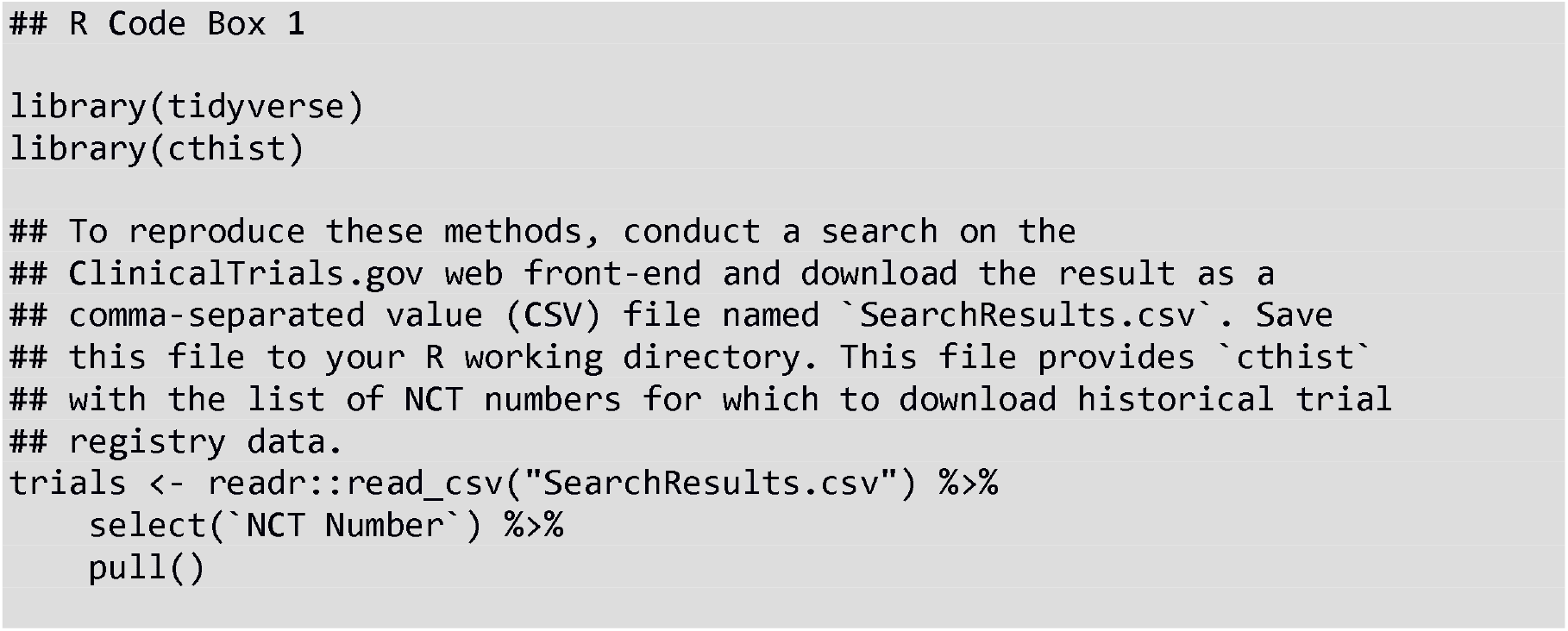
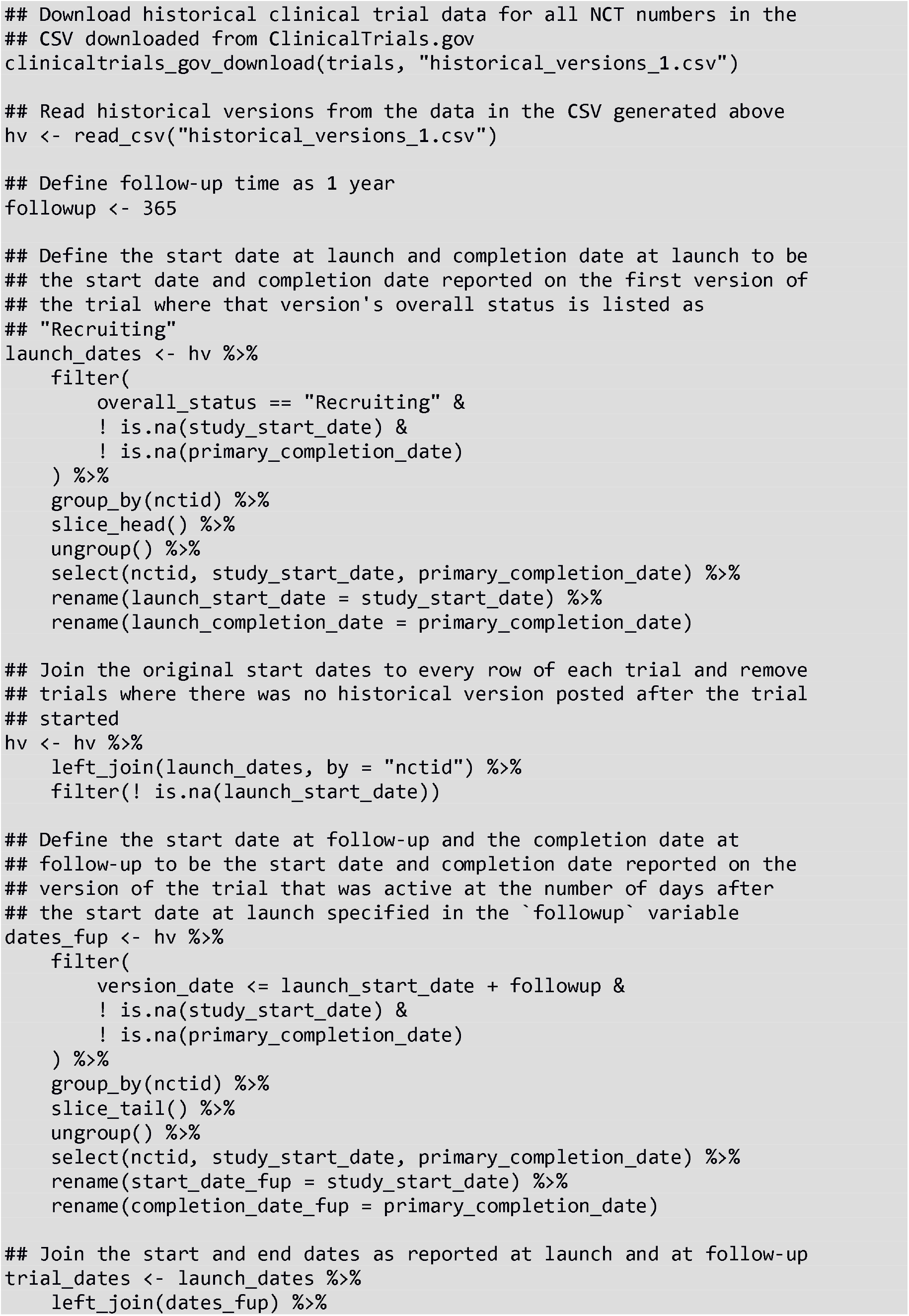
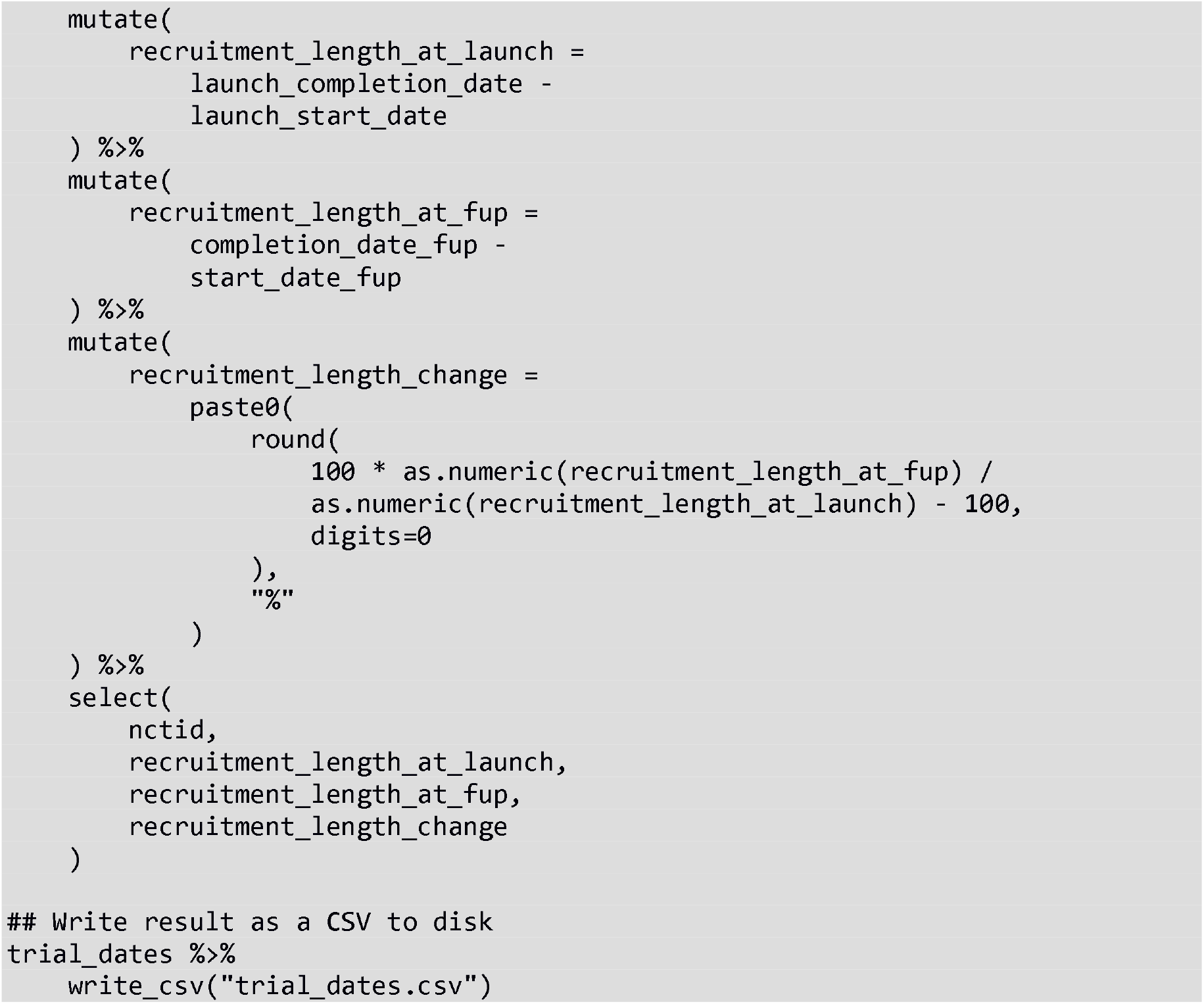

### Case study 2: identifying changes to outcome measures

Outcome switching in clinical trials is a common practice^20^ in which the outcomes that were pre-specified in a clinical trial registry differ from those that are published in the corresponding journal publication. Unreported outcome switching may mislead readers or introduce bias.^21^ Because a clinical trial registry entry may be updated at any time, it is necessary to consult not just the most recent version of a clinical trial registry entry, but to review all the versions in the trial registry history in order to determine whether, when, and in what manner they were changed.

The code presented in *R Code Box 2* will take a list of DRKS id’s from a CSV downloaded from the web front-end of DRKS.de, download all the historical versions of those trials and determine which updates to each clinical trial represents a change in the trial’s outcome measures. This script will identify all changes, from ones as major as the malicious switching of primary and secondary outcomes, to ones as minor as the correction of typos, or even a single-character white-space change. The work of determining whether the change should be mentioned in a final journal publication remains to be done by human curation of the script’s output data.

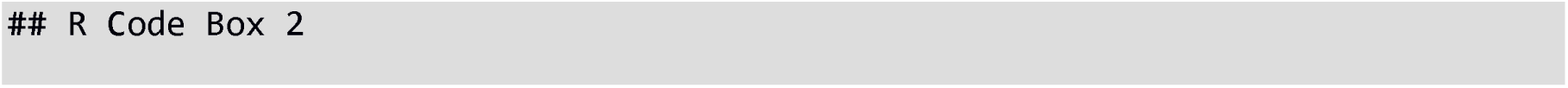
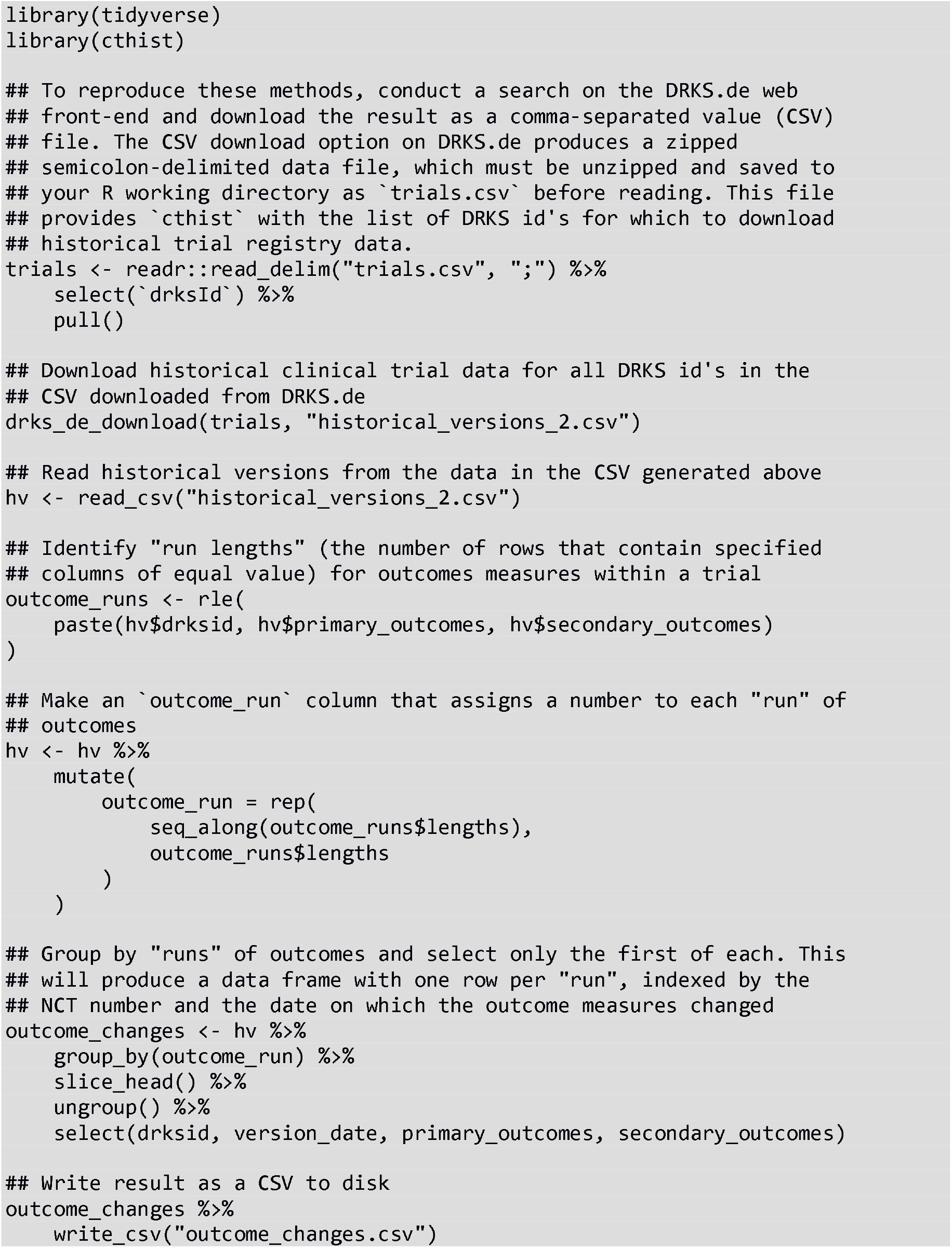

### Case study 3: correcting for variable follow-up time

Let us consider the hypothetical case of a meta-researcher who wishes to characterize phase 3 glioblastoma clinical trial activity in terms of how many clinical trials are stopped (overall status changed to “Terminated”, “Suspended” or “Withdrawn”). It may be tempting to search on the ClinicalTrials.gov web front-end for all or phase 3 trials with an indication of “glioblastoma” whose overall status is “Terminated”, “Suspended” or “Withdrawn”, count the results and report them as a fraction of the number of results for the same search without the overall status restriction. As of the writing of this manuscript, 17.2% phase 3 glioblastoma trials on ClinicalTrials.gov (16 out of 93 on 2022-01-05) had an overall status of “Terminated”, “Suspended” or “Withdrawn”.

This strategy does not account for variable follow-up time among the clinical trials in the sample. A trial that was registered yesterday, for example, may yet go on to be withdrawn, if it were given the same follow-up time as the trials in the sample that were registered five years ago. By failing to account for variable follow-up, our hypothetical researcher is systematically introducing bias into their sample and may produce a misleading count of the number of stopped trials. To correct for this, the overall status of every trial in the sample must be assessed at the same follow-up time; trials where the requisite follow-up time has not yet passed must be excluded from analysis.

The script in *R Code Box 3* will download historical clinical trial data for the trial numbers specified in the CSV downloaded from a search using the web front-end of ClinicalTrials.gov. Trials with less than five years of follow-up will be removed from the sample, and the script will write a CSV file to disk that includes the overall status of each eligible trial at five years after the initial registration.

Among the 93 phase 3 glioblastoma trials on ClinicalTrials.gov as of 2022-01-05, 69 have 5 years of follow-up, and 10 of those (14.4%) had a status of “Terminated”, “Suspended” or “Withdrawn” at 5 years. Among the 6 trials that have overall statuses of “Terminated”, “Suspended” or “Withdrawn” on the most recent version on ClinicalTrials.gov but not at 5-years of follow-up, 4 were originally registered less than 5 years ago, and the remaining 2 changed their status to “Terminated”, “Suspended” or “Withdrawn” after the 5-year mark.

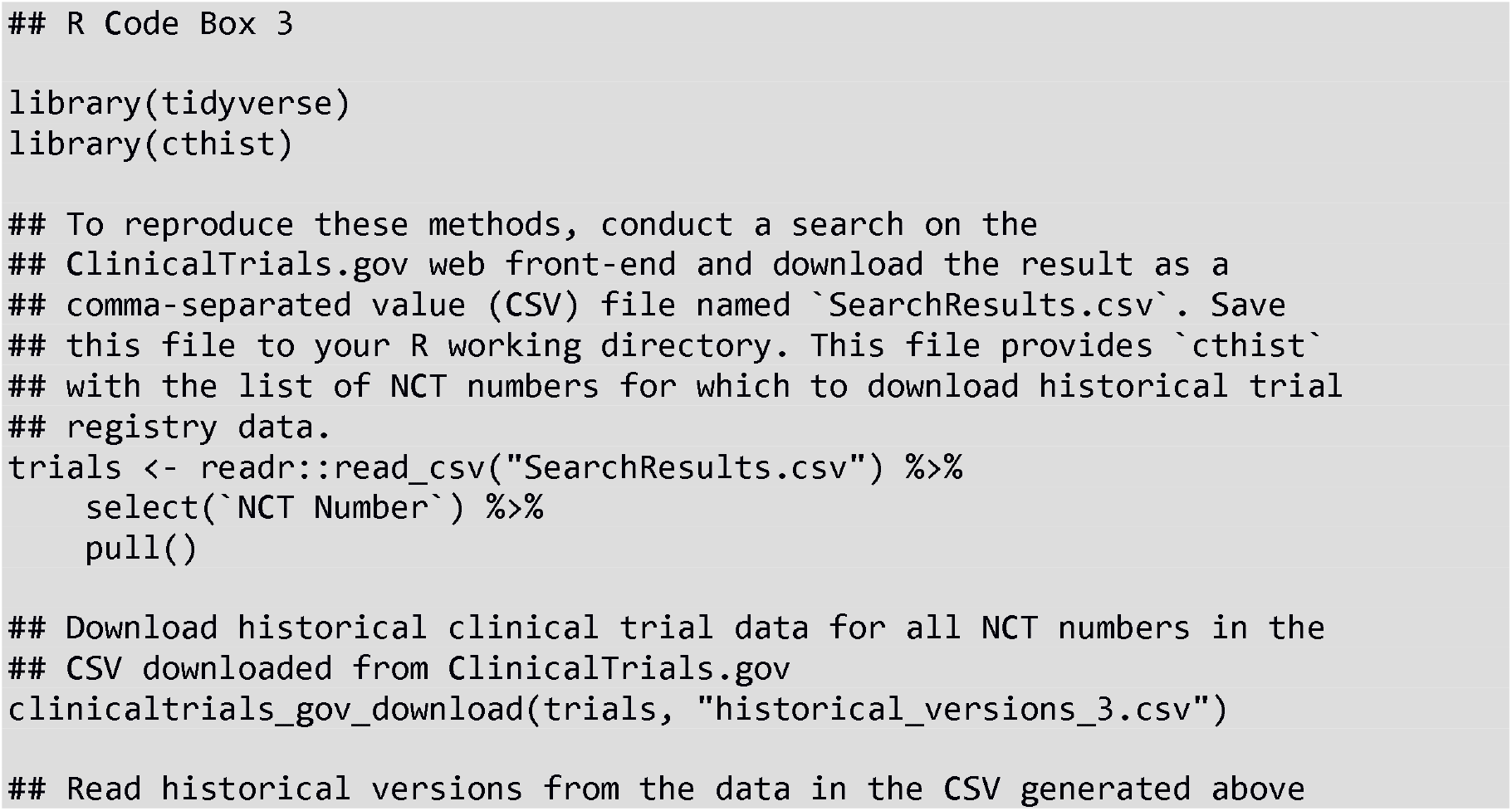
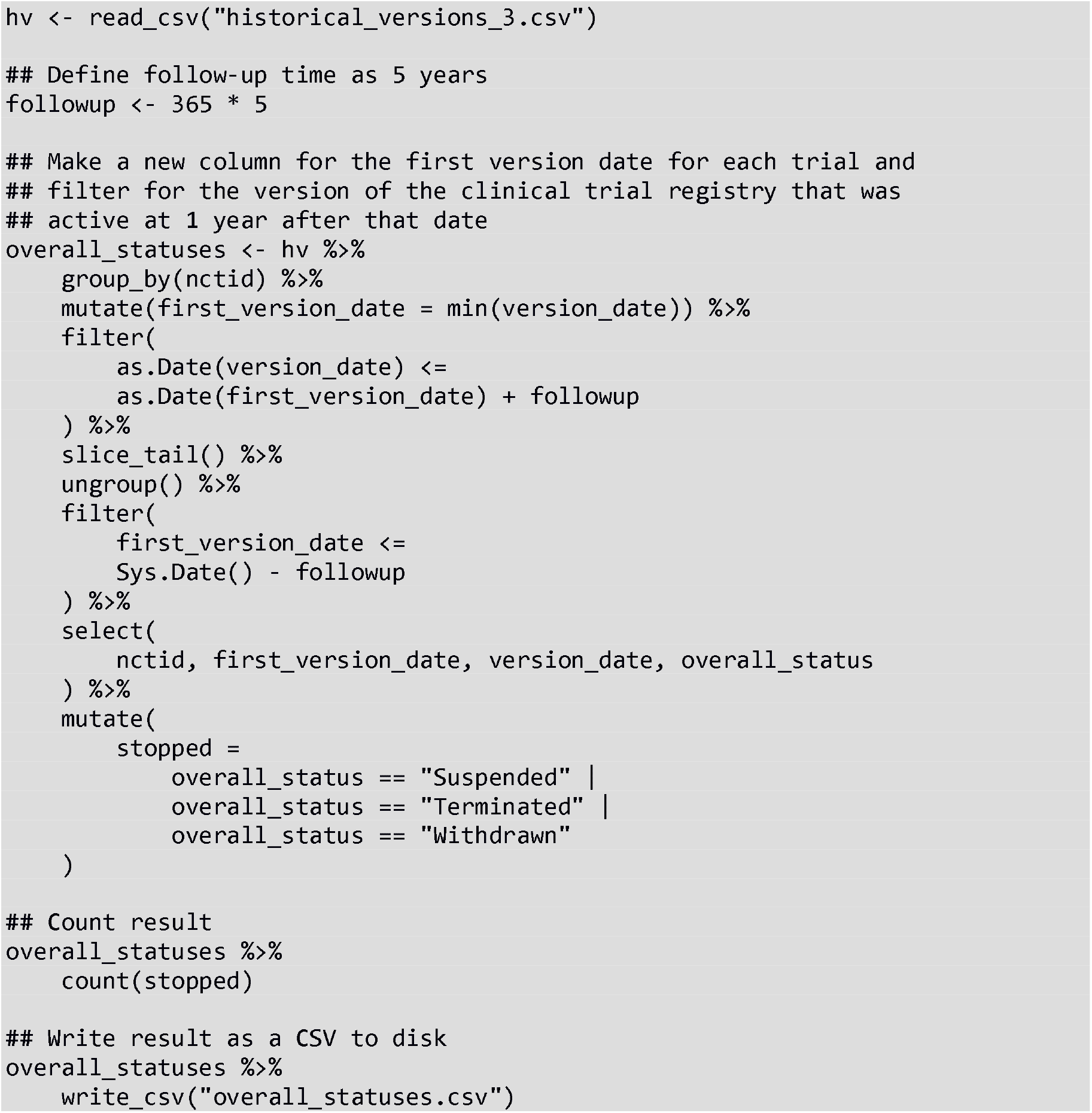

## Discussion

The purpose of prospective registration of clinical trials is partly vitiated if there is no efficient means to access historical clinical trial registry data. The responsible party for a clinical trial can effectively “bury” changes to a registry entry in its history if there is no feasible way to access history changes. Peer-review of individual clinical trial journal publications does not provide sufficient scrutiny to ensure that the publication is not misleading or biased due to outcome switching, etc. as trial registration records are not always thoroughly checked for accuracy.^22^ While the disclosure of changes to individual trial registry entries through the registry website provides some level of openness, this only allows researchers to find registry entry changes if they already know where to look, and limits the feasibility or reproducibility of certain kinds of research.

The `cthist` package provides functions that allow for efficient mass-downloading and processing of historical clinical trial registration data from ClinicalTrials.gov and DRKS.de. This makes certain kinds of meta-research feasible and provides a means to correct common errors in data collection and analysis, such as overlooking variable follow-up. This package also increases the reproducibility of previously completed analyses of clinical trial registry data. Among analyses of clinical trial registry entries, it is common practice to report the date that the clinical trial registry was searched, as the database’s contents change frequently. Without a way to select data from a specific date as provided by `cthist`, reproducing this kind of research is difficult or impossible.

### Limitations

The *R* package described here is a web-scraper that provides a means for retrieving historical clinical trial data that is not easily available without extensive manual work. Web-scraping is less than ideal because a clinical trial registry may change its website at any time in ways that alter CSS selectors on which this web-scraper depends, for example.

Not all data points that are available in ClinicalTrials.gov or DRKS.de are collected by `cthist`, although the package is easily extensible to collect anything reported on the historical version page.

### Future directions

The WHO registry network lists 17 primary clinical trial registries other than ClinicalTrials.gov (including DRKS.de).^23^ Future versions of this package may include functions for downloading historical clinical trial registry data from other clinical trial registries. Future versions may also include functions for downloading additional data points from ClinicalTrials.gov and DRKS.de.

This *R* package may also be integrated into an automated tool to generate reports for reviewers of clinical trials in partnership with journals who publish clinical trials results. This report could include key information on a clinical trial based on data extracted by `cthist` to assist in their reviews. A prototype of such an application is available.^24^

It is also my hope that the existence of this *R* package may draw attention to its necessity by those who make design decisions for clinical trial registries, and implement means for mass-downloading historical clinical trial data for analysis that do not require the use of this *R* package.

Data point ClinicalTrials.gov DRKS.de

## Data Availability

All data produced are available online at https://github.com/bgcarlisle/cthist

## Acknowledgements

This work was informed and motivated by clinical trial research projects involving Delwen Franzen, Martin Haslberger, Martin Holst, Nora Hutchison, Katarzyna Klas, Maia Salholz-Hillel, Daniel Strech and Marcin Waligora.

